# Genomic Modifiers of Neurological Resilience in a Niemann-Pick C family

**DOI:** 10.1101/2025.03.03.25321546

**Authors:** Macarena Las Heras, Benjamín Szenfeld, Valeria Olguín, Juan Carlos Rubilar, Juan Francisco Calderón, Yanireth Jimenez, Silvana Zanlungo, Emanuele Buratti, Andrea Dardis, Francisco A. Cubillos, Andrés D Klein

## Abstract

Niemann-Pick type C (NPC) disease, caused by pathogenic variants in the *NPC1* or *NPC2* genes, disrupts cellular cholesterol and glycolipids trafficking. Patients exhibit a wide spectrum of visceral and neurological manifestations, suggesting a role for genomic modifiers. To uncover the genetic basis of NPC neurological resilience, we analyzed the exomes of an NPC family with diverse phenotypes, from very mild to severe neurological involvement. Linkage analysis revealed loss-of-function (LOF) variants in *CCDC115*, *SLC4A5*, *DEPDC5*, *ETFDH*, *SNRNP200*, and *DOCK1* that co-segregated with resistance to severe neurological signs. Biomarkers of severity are lacking in NPC. Based on LOF variants in the yeast orthologs of these genes, we successfully predicted NPC-like severity in *Saccharomyces cerevisiae* of different genetic backgrounds. Complementary, to associate pathways with severity, we performed RNA-seq, uncovering positive correlations between mitochondrial transcripts with cellular fitness. Finally, we modeled NPC disease in yeast lacking the sodium bicarbonate cotransporter *bor1*, the *SLC4A5* ortholog. Deletion of *bor1* enhanced cellular fitness, prevented vacuolar fragmentation, reduced sterols buildup, and improved mitochondrial function. Our study revealed modifiers/biomarkers of NPC severity, and highlighted *SLC4A5* as a promising therapeutic target for this devastating disease.

## Introduction

Niemann-Pick type C (NPC) disease is a progressive neurovisceral lysosomal storage disorder resulting from loss-of-function pathogenic variants in the *NPC1* or *NPC2* genes, leading to abnormal cholesterol and lipid accumulation within lysosomes (Klein et al., 2014; Platt, 2014). Disease onset and severity of manifestations exhibit significant variability, even within the same family, highlighting the influence of genetic and/or non-genetic factors, known as disease modifiers (Bolton et al., 2022; Las Heras et al., 2023; Wassif et al., 2016).

Understanding the impact of genomic modifiers, *loci* that enhance or suppress the effects of a primary disease-causing variants (Nadeau, 2001, 2003), is crucial for elucidating the mechanisms underlying the diverse clinical presentations of NPC (Las Heras et al., 2023). From a translational perspective, genomic modifiers hold promise as disease biomarkers and therapeutic targets (Cahill et al., 2022).

Common approaches to map modifier genes in monogenic diseases include Genome-Wide Association Studies (GWAS) and linkage analysis (Génin et al., 2008; Riordan & Nadeau, 2017). When samples derived from patients with different severities are not available, modeling diseases in organisms of multiple genetic backgrounds can facilitate the discovery and validation process (Olguín et al., 2022). Recent studies in neurodegenerative proteinopathies such as Alzheimeŕs, Parkinson’s, polyglutamine expansion diseases, and in lysosomal storage disorders (LSDs) have identified genomic modifiers using model organisms, including mice, flies, worms, and yeast, emphasizing the critical role of genetic background in shaping disease phenotypes (Farhat et al., 2024; Klein et al., 2016; Na et al., 2013). However, research on NPC modifiers remains limited, partly due to the scarcity of extensive, annotated genomic data from patients with well-characterized phenotypes suitable for GWAS studies. In this sense, model organisms can complement patient-based discovery and validations.

Our study combined human linkage analysis with a yeast model to investigate the role of genomic modifiers in NPC disease, providing a novel approach to identifying biomarkers and potential therapeutic targets. We conducted a linkage analysis within an NPC family exhibiting diverse clinical manifestations, ranging from very mild to severe neurological symptoms. The analysis revealed co-segregation of loss-of-function (LOF) variants in six genes among NPC individuals protected for early neurological symptoms. We utilized variants in their yeast orthologs to successfully predict NPC severity in *Saccharomyces cerevisiae* of different genetic backgrounds treated with U18666A (U18), a chemical inhibitor of Ncr1p, the functional and structural ortholog of the human NPC1 protein. We characterized the gene expression patterns of the NPC-like yeast and observed a correlation between mitochondrial transcripts and cellular fitness. Finally, we validated that *SLC4A5*, a gene identified in the human linkage analysis, has a modifying role in the NPC-like phenotype. The U18-treated Δ*bor1* yeast presented improved cellular fitness, decreased vacuolar fragmentation, reduced filipin fluorescence, and improved mitochondrial function assessed by NAD/NADH ratio, compared to U18-treated wild-type yeast, highlighting its potential as a therapeutic target.

## Materials and Methods

### Exome sequencing and variant calling

We performed exome sequencing of 11 members belonging to an NPC family followed at the Regional coordinator Centre for Rare Disease, University Hospital of Udine. Five of them had been diagnosed as affected by the disease based on positive filipin staining and the identification of two missense variant in compound heterozygosis in the *NPC1* gene: c.1351G>A, p.(E451K) and c.2974G>T, p.(G992W). Phenotypically, affected siblings were classified according to the age at onset of neurological symptoms (Geberhiwot et al., 2018). Patients who did not present clinical or imaging signs of neurological involvement at last follow up were considered non-classifiable (NC). The study was authorized by the Regional Ethics Committee (66/2014/OS; data 18/11/2014) and conducted according to the declaration of Helsinki. Signed informed consent was collected from patients (or their parents in case of minors). Genomic DNA sequencing was conducted at the EMBL Genomics Core facility using DNBseq™ next-generation sequencing platform and the Agilent SureSelect Human All Exon V6+UTR library kit, achieving an average coverage depth of 100×.

Variant analysis followed the best practice recommendations of the Genome Analysis Toolkit (GATK) (DePristo et al., 2011). Briefly, raw sequencing reads were aligned to the human reference genome (hg19) using the Burrows–Wheeler Aligner (BWA) (Li & Durbin, 2009), generating BAM files. PCR duplicates were removed using Picard tools (http://broadinstitute.github.io/picard) to mitigate biases from PCR amplification. Base quality score recalibration and local realignment around indels were performed using GATK’s BaseRecalibrator and IndelRealigner tools, respectively (McKenna et al., 2010), to improve the accuracy of variant calling. Variants were called using GATK’s HaplotypeCaller, and variant quality scores were recalibrated using GATK’s VariantRecalibrator to generate a Variant Quality Score Logarithm of Odds (VQSLOD) (McKenna et al., 2010).

### Linkage Analysis Using Human Data (VAAST Analysis)

To identify candidate modifier genes, gene-based variant association analyses were performed using the Variant Annotation, Analysis, and Search Tool (VAAST) (Hu et al., 2013). VAAST 2.0 uses an aggregative variant association test that combines parameters such as amino acid substitutions, allele frequencies, and phylogenetic conservation to rank genes based on the presence of potentially functional variants that differ between cohorts.

These analyses utilized both genotypic and phenotypic data. Genotypic data consisted of filtered variants, while phenotypic data included detailed clinical information from affected members of NPC families exhibiting varying levels of disease severity. Patients were categorized into predefined phenotypic subgroups: presence or absence of neurological, psychiatric, and visceral symptoms. Subsequently, the VAAST 2.0 algorithm was run under both dominant and recessive patterns to evaluate associations with each phenotypic group. The resulting gene rankings were generated based on VAAST 2.0 probability scores.

### SIFT and PolyPhen-2 for predicting variant pathogenicity

Variants identified in the top-ranking genes of each linkage were further assessed using *in silico* pathogenicity prediction algorithms, including Sorting Intolerant From Tolerant (SIFT) (Kumar et al., 2009) and PolyPhen-2 (I. Adzhubei et al., 2013). For predicting the role of each variant in *S. Cerevisiae* we aligned the protein sequences of all strains using MUSCLE, a multiple-sequence alignment tool (Edgar, 2004), within the Geneious software. SIFT predictions were conducted via the SIFT web server (https://sift.bii.a-star.edu.sg), where the identified substitutions were evaluated based on sequence homology and amino acid physicochemical properties. Variants were classified as either “tolerated” or “deleterious,” with a SIFT score ≤ 0.05 indicating a deleterious effect. Similarly, functional predictions were obtained using the PolyPhen-2 web server (http://genetics.bwh.harvard.edu/pph2/). PolyPhen-2 classified variants as “benign,” “possibly damaging,” or “probably damaging” by integrating structural modeling, sequence conservation, and physicochemical properties.

### Identification of orthologs of the putative modifiers

To identify *Saccharomyces cerevisiae* orthologs of human genes, curated orthology annotations from the Saccharomyces Genome Database (SGD) and the Ensembl Genome Browser were employed.

In SGD, orthology information was obtained from the “Comparative Info” section of each gene entry. Orthologs relationships in SGD are established based on comparative genomic analyses integrating manually curated sources, including literature-supported functional studies and computational predictions. Additionally, SGD incorporates orthology assignments from large-scale comparative genomic studies such as the Yeast Gene Order Browser (YGOB) and curated homologous relationships from model organisms.

In Ensembl, orthology data were retrieved from the “Comparative Genomics” > “Orthologs”, where orthologs relationships are inferred using phylogenetic gene tree reconstruction within the Ensembl Compara pipeline. This pipeline integrates multiple sequence alignments, phylogenetic inference, and synteny conservation to classify orthologs into one-to-one, one-to-many, or many-to-many relationships, providing a robust framework for evolutionary comparisons across species.

### Yeast strains

The following strains were used: Y12 [Sake, (“SA”): Mat alpha *ho*::NatMX, *ura3*::KanMX], YPS128 [North American (“NA”): Mat alpha *ho*::NatMX, *ura3*::KanMX], DBVPG6044 [West African, (“WA”): Mat alpha *ho*:: NatMX, *ura3*::KanMX] and DBVPG6765 [Wine/European, (“WE”): Mat alpha *ho*::NatMX, *ura3*::KanMX]. These strains were generously provided by Dr. Francisco Cubillos (Cubillos et al., 2009). Validation studies were performed using the BY4742 strain, specifically the wild-type (WT; Accession Y10000) and the *bor1*-deletant (Δ*bor1*; Accession Y11169). Both strains were obtained from Euroscarf (http://www.euroscarf.de).

### Cell growth

Yeast were precultured in 200 µL of YNB medium (0.67% YNB without amino acids supplemented with 2% glucose, 0.2% uracil, and with 0.0875% yeast synthetic drop-out medium without uracil) for 48 h at 28°C. Subsequently, the yeast were inoculated to an optical density (OD) of 0.03-0.1 (wavelength 620 nm) in 200 µL of medium (with and without 200 μg/mL U18666A) and incubated without stirring at 25°C for 36-48 h in a Tecan Sunrise absorbance microplate reader. The OD was measured every 30 minutes using a 620 nm filter. Each experiment was performed in triplicate. Maximum growth rates (μMax), lag time, and OD max parameters were obtained using GrowthRates software, which adjusts the OD versus time measurements to the re-parameterized Gompertz equation proposed by Zwietering (Zwietering et al., 1990).

### RNA-sequencing

Transcriptomic analyses of the four yeast strains under control and U18-treatments were performed in three biological replicates. For this, yeast taken from patches on YPD plates were grown in triplicates at 28 °C until the mid-log phase (OD600∼0.8) under two conditions: control (5 mL of YNB medium) and treated with U18 (5 mL of YNB medium containing 200 µg/mL U18666A). The cells were then washed twice with 1 mL of 50 mM EDTA, centrifuged at 5000 rpm for 3 minutes each time, and resuspended in 1 mL of solution SORBITOL 0.1M, EDTA 0.1M, pH 7.4 (solution Y1) which contained beta-mercaptoethanol (2-ME) and 2 U of zymolase. The suspension was incubated for 30 minutes at 37°C. After incubation, the cells were centrifuged at 1000 rpm for 5 minutes, and the supernatant was carefully discarded to avoid loss of the pellet. RNA was extracted using the E.Z.N.A. Total RNA Kit I (OMEGA) according to the manufacturer’s instructions. To eliminate genomic DNA contamination, the samples were treated with DNase I (Promega). RNA was then cleaned and concentrated using the Thermo GeneJET RNA CleanUp and Concentration Kit, following the manufacturer’s protocol. RNA integrity was assessed using a Fragment Analyzer (Agilent).

The samples were then sent to BGI, where the preparation of the libraries and sequencing were carried out employing DNBseq^TM^ technology. The RNA-seq data was deposited in the Gene Expression Omnibus (GEO) database, www.ncbi.nlm.nih.gov/geo (accession no GSE286511).

### RNA-Seq analysis

RNA-Seq data were analyzed using the 3D RNA-seq platform (https://3drnaseq.hutton.ac.uk/app_direct/3DRNAseq/), an interactive R-based tool that facilitates comprehensive transcriptomic analysis and employs the limma statistical framework for accurate differential expression analysis (Ritchie et al., 2015).

Raw RNA-Seq FASTQ files were first uploaded to Galaxy Europe, where paired datasets (forward and reverse reads) were generated for each sample. Sequence mapping and transcript quantification were performed using the Salmon algorithm (Patro et al., 2017), which aligns reads to the reference transcriptome and quantifies gene expression in terms of transcripts per million (TPM) and raw counts for subsequent analyses. Differentially expressed genes were identified across experimental conditions and genetic backgrounds using the 3D RNA-seq platform.

Subsequently, functional enrichment analyses were performed to identify biological pathways and processes associated with the transcriptional changes observed across the strains, aiming to uncover key genes and regulatory pathways contributing to phenotypic differences. Pathway enrichment analysis was conducted using the Kyoto Encyclopedia of Genes and Genomes (KEGG) database with the clusterProfiler package in R, employing the enrichKEGG function and specifying *Saccharomyces cerevisiae* (sce) as the organism. Gene Ontology (GO) enrichment analysis was performed separately for the three GO ontologies: Biological Process (BP), Cellular Component (CC), and Molecular Function (MF). BP describes the biological processes involving the genes, CC specifies the cellular localization of the genes, and MF focuses on the biochemical activities of the genes. Overrepresented GO terms were identified in each analysis, and p-values were calculated to assess their statistical significance. The enrichGO function, utilizing the org.Sc.sgd.db package, was employed for these analyses, specifying “ORF” as the key type. Visualization of KEGG and GO results was performed using the enrichplot package, with dot plots generated to facilitate interpretation of enriched pathways and terms. Additional R packages, such as ggplot2, dplyr, openxlsx, extrafont, stringr, and scales, were employed for data manipulation and visualization. Together, these analyses provided a comprehensive understanding of the biological processes, cellular localizations, and molecular functions underlying the observed transcriptional and phenotypic variation across strains.

### FM^TM^ 4-64 staining and vacuolar morphology quantification

FM dyes are lipophilic styryl compounds commonly employed in diverse studies focusing on plasma membrane and vesicle dynamics. Here, FM^TM^ 4-64 FX (Thermo Fisher, Cat. no. F34653) was utilized to stain yeast vacuolar membranes (excitation/emission maxima ∼565/744 nm) and classify vacuolar morphologies into classes “A”, “B” or “C”, according to the criteria established by Seeley and colleagues (Seeley et al., 2002). Briefly, yeast cells were taken from patches on YPD-agar plates and inoculated into 500 µL of YPD medium, with and without 200 µg/mL U18666A, containing 3 mM FM^TM^ 4-64 FX. The cultures were grown for 24 hours at 28°C with constant shaking. Samples of 4 µL were examined using a Leica SP8 confocal microscope (Leica Microsystems, Germany). A total of 110-150 cells were analyzed per condition, and their vacuolar morphologies were categorized. The experiments were performed in triplicate.

### Filipin staining

Filipin staining was used to evaluate the ergosterol buildup in yeast (the yeast equivalent of cholesterol), as previously described (Van Leeuwen et al., 2008). Briefly, yeast were taken from patches on YPD plates and inoculated into 500 µL of YPD (with and without 200 µg/mL U18666A). The cultures were grown for 24 hours at 28°C with constant shaking. Then, 7 ml of 2.14 mM filipin was added and incubated for 15 minutes. Samples of 3 µL were examined using a fluorescence microscope (excitation at 340-380 nm and emission at 385-470 nm).

### NAD^+^/NADH Assay

Intracellular NAD^+^ and NADH levels and NAD^+^/NADH ratio were determined by NAD^+^/NADH Assay Kit (Abcam, Cat #65348) according to manufacturer’s instructions, with modifications. Briefly, 3 mL of yeast culture per condition was harvested, washed with ice-cold PBS, and lysed using 400 μL of extraction buffer through three cycles of freezing (liquid nitrogen), vortex and thawing. The lysate was centrifuged at 10,000 g for 10 min at 4°C to remove proteins. Then, half of supernatant volume was incubated at 60°C for 30 min to decompose NAD^+^, while the other half volume was designated as total NAD (NADH plus NAD^+^). 1.25 μL of each half of the supernatant was then transferred to a clear-bottom 96-well plate. For each assay, a series of NADH standards of 0, 10, 20, 30, 40, and 50 pmol/well, were included. 50 μL of NAD cycling buffer and enzyme mix (49 μL cycling buffer and 1 μL cycling enzyme mix from the manufacture) was added to each sample and incubated for 5 min at room temperature to convert all NAD^+^ to NADH. 5 μL of manufacture’s NADH developer was added into each well. Absorbance at 450 nm was measured using a plate reader after 2 hours of incubation. Total NADt and NADH levels were calculated using a standard curve and expressed as pmol/OD.

### Statistics

We used Prism v9.5.0 for statistical analyses (GraphPad Software, San Diego, CA). At least three biological replicates were studied under experimental conditions. Data are presented as the mean value ± standard error of the mean (SEM). We performed the Shapiro-Wilk normality test (p > 0.05) to assess data distribution. For normally distributed data, we conducted one-way ANOVA followed by Tukey’s post-test (p < 0.05) with multiple comparisons. For non-normally distributed data, we used the Kruskal-Wallis test (p < 0.05), followed by multiple comparison tests when appropriate. The specific statistical tests used are indicated in the corresponding figure legends. A p-value < 0.05 was considered statistically significant.

## Results

### An NPC family showing a broad spectrum of phenotypic variability

We studied an NPC family with five members siblings affected by NPC carrying the with a c.1351G>A, p.(E451K) and c.2974G>T, p.(G992W) variants in the *NPC1* gene in compound heterozygosis. The clinical presentation varied greatly among the affected individuals in terms of disease severity ranging from mild visceral disease to severe neurological and psychiatric involvement (Table S1). The broad phenotypic spectrum observed in this family provides a unique opportunity to explore genetic modifiers underlying the heterogeneity in symptom presentation.

### Linkage analysis identifies putative modifier genes of NPC severity

Linkage analysis can identify symptom-causing genes with large effect sizes by searching for co-segregation of variants with a phenotype within a family. To identify potential modifier genes influencing NPC severity disease, we conducted multiple linkage analyses on the same family (Figure 1, Table S2). We categorized individuals based on phenotypic classification, as juvenile vs adult/NC siblings.

**Figure 1.**
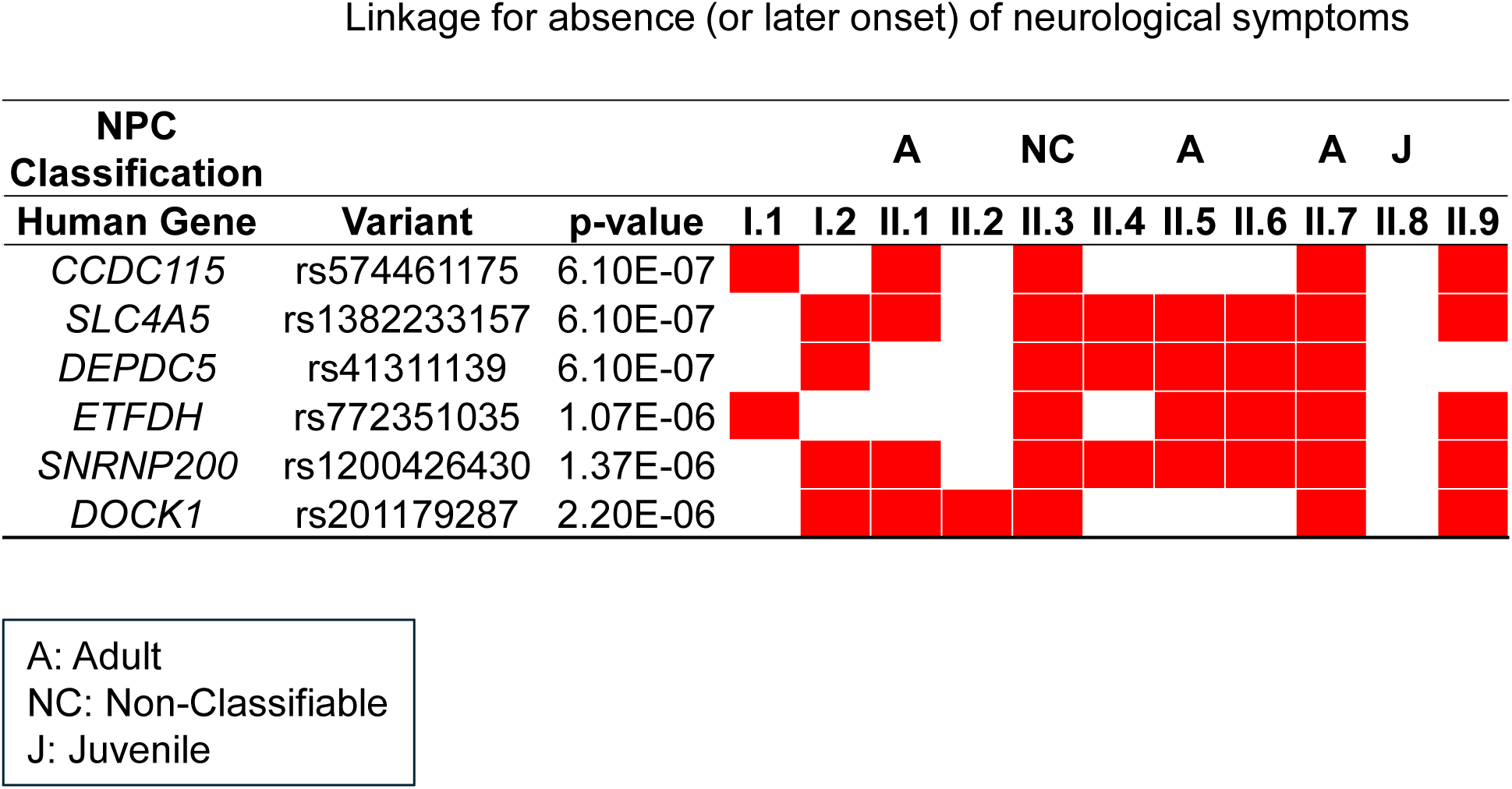
Linkage analysis of an NPC family with phenotypic variability. Diagram of linkage analysis for absence or latter onset of neurological symptoms, including gene names, variants, p values of the linkage, and the distribution of loss-of-function (LOF) variants (in red) among family members. The NPC classification of the individuals is also included (A: Adult; NC: Non-Classifiable; J: Juvenile).

We employed VAAST 2.0 algorithm for linkage because it assesses the cumulative impact of both coding and non-coding variants, prioritizes candidate genes by considering both dominant and recessive inheritance patterns, and has been used to map modifiers (Hu et al., 2013; Jimenez et al., 2022; Yandell et al., 2011). VAAST 2.0 analysis identified numerous genes with a Composite Likelihood Ratio Test (CLRT)-derived p-value < 0.05 in each analysis. Following Bonferroni correction for multiple comparisons, the significance threshold was adjusted to 2.48E-06. For example, the analysis identified 36 genes associated with the presence of neurological symptoms under a dominant inheritance pattern and 30 genes under a recessive inheritance pattern. One of them, *COPZ2*, has been implicated in intracellular protein trafficking and is linked to Alzheimer’s disease (Yang et al., 2019). These results represent a subset of the broader analysis. The comprehensive lists of statistically significant genes identified in each analysis are provided in Supplementary Table S2.

Given our focus on translational research, we prioritized the linkage analysis for protection from early neurological symptoms. Among the identified genes, those with predicted LOF variants are particularly promising therapeutic targets because inhibiting a gene function is generally more feasible than enhancing it (Minikel et al., 2020). Usually, variants identified by linkage analysis have a large size effect in pathological cascades, serving as well as valuable genomic biomarkers.

To evaluate the effects of the amino acid substitutions in each strain, we employed SIFT (Sorting Intolerant From Tolerant) (Ng & Henikoff, 2003; Sim et al., 2012) and PolyPhen-2 (Polymorphism Phenotyping v2) (I. A. Adzhubei et al., 2010), two widely used computational tools for predicting the functional consequences of missense variants. We identified heterozygous deleterious variants within *CCDC115* (p=6.1E-07), *SLC4A5* (p=6.1E-07), *DEPDC5* (p=6.1E-07), *ETFDH* (p=1.0E-07), *SNRNP200* (p=1.3E-06), and *DOCK1* (p=2.2E-06) genes (Figure 1). These variants were present in individuals II.1, II.3, II.5, and II.7, but absent in individual II.8, who exhibited a juvenile neurological phenotype characterized by early onset and significant symptom severity (Baxter et al., 2022; Patterson et al., 2017; Vanier, 2010). We hypothesized that these variants may serve as biomarkers for predicting severity and as therapeutic targets.

### Genomic modifiers as disease severity biomarkers in NPC-like yeast of different genetic backgrounds

The study of a single NPC family served as an identification cohort. However, replicating these findings in a separate cohort is crucial to validate their applicability as biomarkers. Due to the lack of accessible genomic data from NPC patients with comprehensive clinical information, we harnessed the genomic variability of Sake (SA), West African (WA), North American (NA), and West European (WE) haploid yeast strains, because their different phylogenic origins and their genomes are fully sequenced (Bergström et al., 2014; Cubillos et al., 2013; Liti et al., 2009). We searched for amino acid substitutions within the yeast orthologs of *CCDC115*, *SLC4A5*, *DEPDC5*, *ETFDH*, *SNRNP200*, and *DOCK1* across the different strains (Figure 2A, Table S3). In keeping with our approach of focusing primarily on LOF variants, we used SIFT and PolyPhen-2 for uncovering LOF variants in yeast as well. Each variant identified as potentially deleterious was assigned a score of one. Subsequently, the scores were summed to determine the overall count of deleterious variants within the orthologs of putative neuroprotective genes for each yeast strain. We did not find LOF variants in the yeast ortholog of *CCDC115* across the strains. Thus, we used the genomic variants within the five remaining yeast orthologs to predict NPC-like severity in yeast. The analysis showed that WA and SA strains had the highest accumulation of such variants in the putative neuroprotective genes. This finding led us to hypothesize that these strains might show greater physiological resistance upon NPC induction with the U18 drug, compared to the NA, and WE strains (Figure 2B).

**Figure 2.**
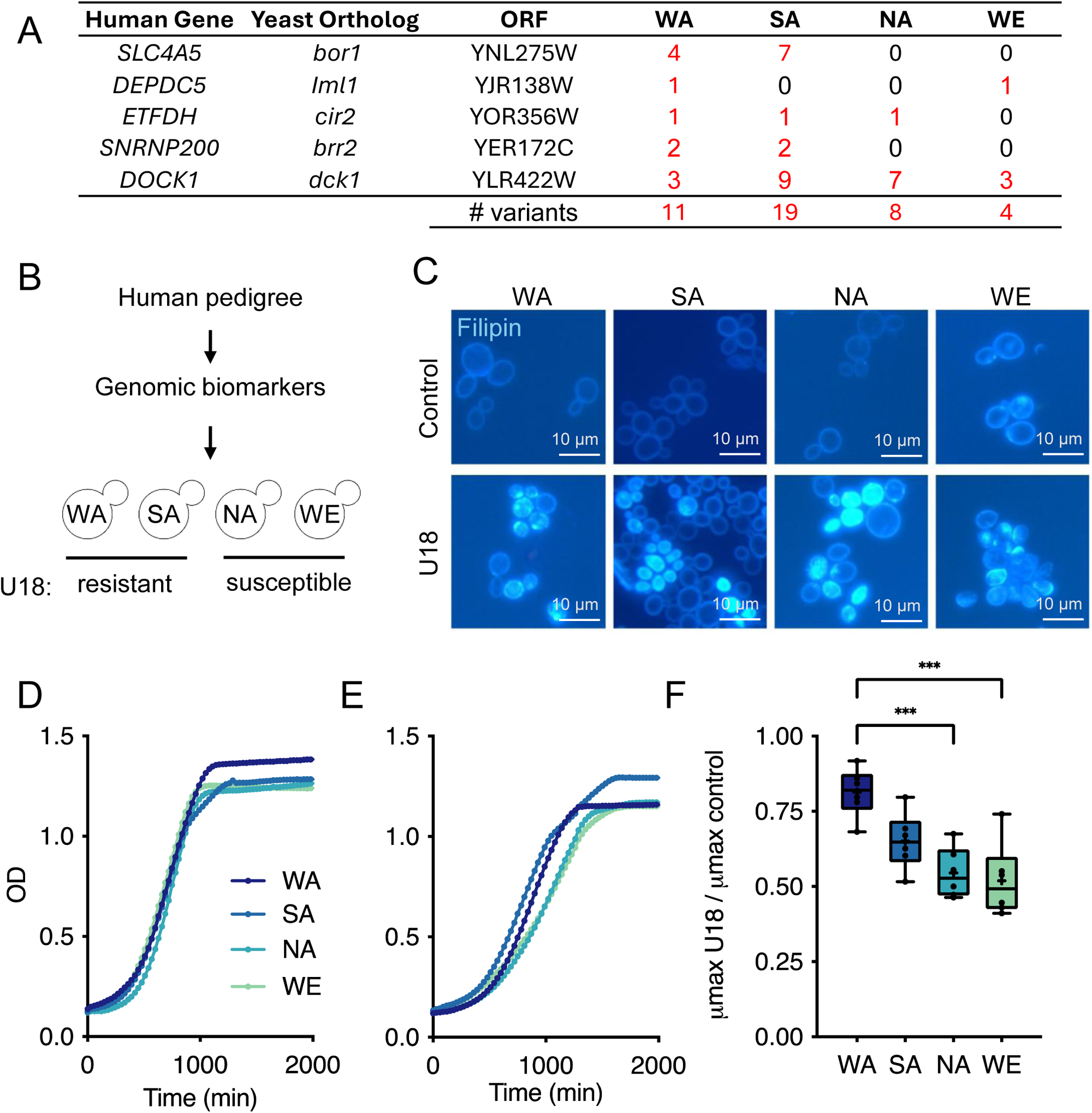
Validation of genomic biomarkers of severity in NPC-like yeast. (A) Summary of the number of LOF variants in the genes identified in the human linkage in each yeast strain. LOF variants are highlighted in red. (B) Predictions of NPC severity in yeast of different genetic backgrounds. (C) Representative images of filipin staining of control and U18-treated (200 μg/mL for 24 h) WA, SA, NA and WE strains. Bars correspond to 10 µM. (D) Growth curves of WA, SA, NA and WE strains (untreated cells). (E) Growth curves of WA, SA, NA and WE strains U18-treated (200 μg/mL), n=7. (F) μMax ratios calculated by dividing the average growth rate of each strain (WA, SA, NA, and WE) treated with U18 by its average growth rate under control conditions (U18/Ctrl). One-way ANOVA and Tukey’s post-test were performed. *p < 0.05, **p < 0.01, ***p < 0.001, ****p < 0.0001. The bars represent the mean ± SEM.

To experimentally validate the predicted severities, we treated the WA, SA, NA, and WE yeast strains with the U18 drug (200 μg/mL for 24h). A hallmark of NPC disease is intracellular cholesterol accumulation (ergosterol in yeast), which can be detected with filipin, a fluorescent probe (Brett et al., 2011; Vanier et al., 1991; Vanier & Latour, 2015). Untreated cells exhibited filipin staining exclusively on the cell membrane. In contrast, U18-treatments induced intense intracellular filipin staining in every strain (Figure 2C), similarly to what is observed in the genetic NPC yeast model (Berger et al., 2005). These NPC-like models were characterized at physiological and transcriptomic levels for biomarker validation and identification.

Growth curves are an essential tool for assessing physiological changes and comparing responses across different conditions. To evaluate the effects of U18, we analyzed the growth curves for each strain, both untreated (control) and after 24 hours of exposure to U18 at 200 μg/mL (Figure 2D-E). From these data, we obtained growth rates (µMax) and calculated ratios by dividing the average growth rate of each strain treated with U18 by the average growth rate in control conditions (Figure 2F). As predicted, the strains showed varied responses to U18 treatment: WA and SA were the most resistant, while NA and WE were the most susceptible. This variability in yeast fitness replicates the phenotypic variability observed in the NPC family and supports the hypothesis that LOF variants in *SLC4A5*, *DEPDC5*, *ETFDH*, *SNRNP200*, and *DOCK1* genes may serve as genomic biomarkers of severity in humans.

### Transcriptional patterns across the NPC-like strains of different genetic backgrounds

Because changes in gene expression might explain the phenotypic differences and uncover additional disease severity biomarkers, we performed RNA sequencing (RNA-Seq) on the four yeast strains, both under control and U18 conditions (Figure 3). The heatmap includes 2,261 significantly expressed genes between U18 and their respective control strain with a 1.5-fold-change threshold and a p≤0.05. The volcano plots show striking differences across the strains (Fig 3A-E). The SA strain, one of the U18 resistant strains, showed little change compared to controls (Fig 3C). The WE strain, U18 susceptible, presented the most different transcriptional patterns (Fig 3A and 3E). Then, we analyzed if the linkage-significant genes were differentially expressed. We did not find significant correlations between µMax and the genes identified in the linkage analysis across the four strains (data not shown). However, we observed a significant reduction in *ETFDH/cir2* and *DOCK1/dck1* in U18-treated WE compared to its control (fold change -1.7 p≤ 0.005 and fold change -1.66 p≤ 0.002, respectively).

**Figure 3.**
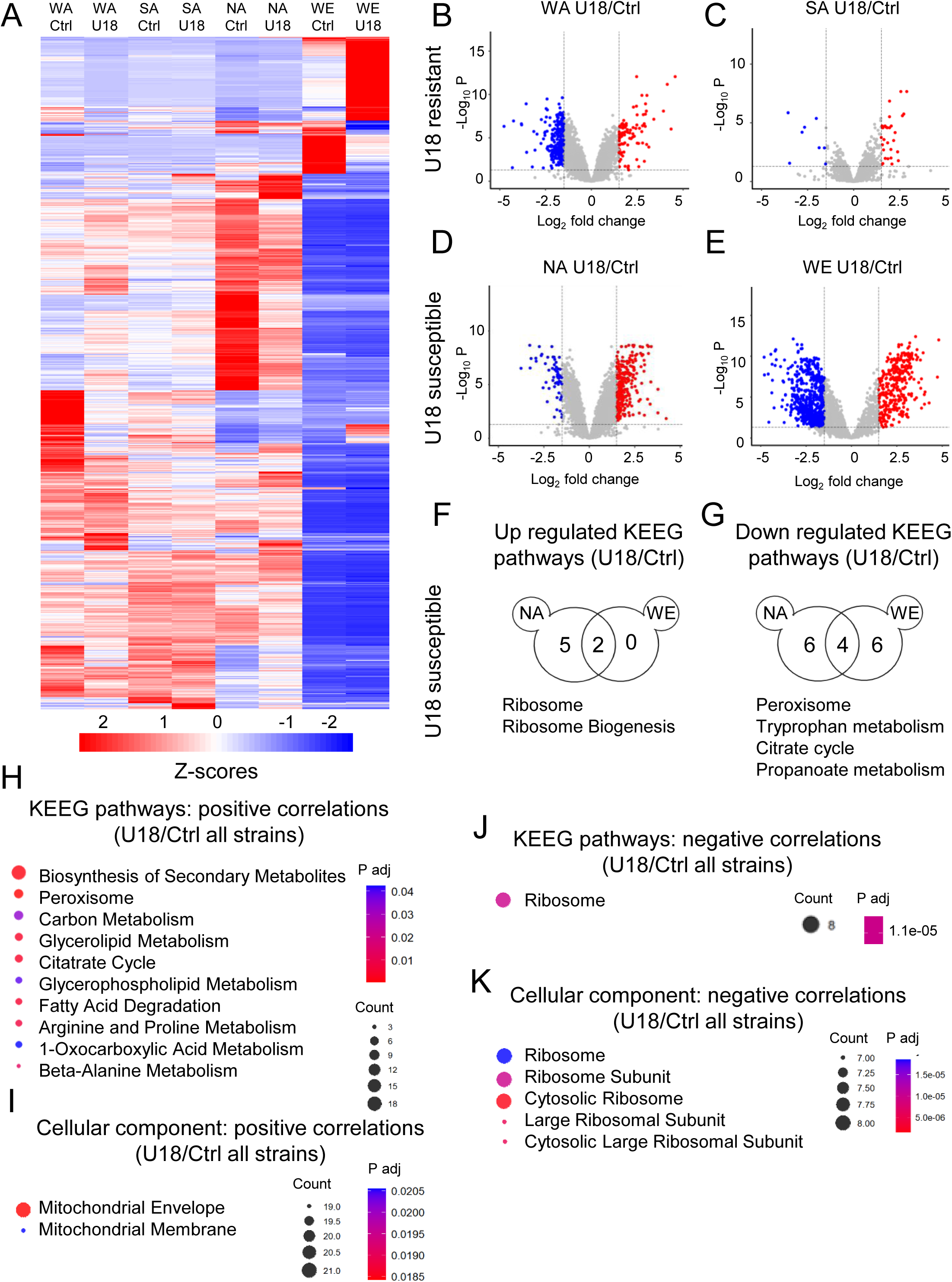
Transcriptomic analysis in the NPC-like yeast. (A) Heatmap of the U18-differentially expressed genes in WA, NA, SA, and WE yeast strains compared to their control strains. (B-E) Normalized volcano plots for each strain (U18/Ctrl). A 1.5-fold-change in expression levels was used as threshold and a p≤0.05. Red and blue dots indicate significantly expressed genes post-treatment. (F-G) Common upregulated and Downregulated KEEG pathways between NA and WE (susceptible) strains. The Venn diagrams indicate the number of significant modulated pathways between the strains. The names of the shared pathways at the intersection of the diagrams are highlighted below. (H-K) The KEEG pathways and Cellular Components correlating with normalized μMax (U18/Ctrl) across all strains are shown. The size of the circle represents the number of counts and the color the p value.

Then we studied if the U18/Ctrl transcriptional patterns were clustered in pathways (Fig 3F-K, Fig S1-4). Initially, we checked for common modulated KEGG pathways between SA and WA (U18-resistant strains), and the NA and WE (U18-susceptible strains). We did not find shared modulated pathways between the resistant, but we did across the susceptible (Fig 3F-G). Among them, the upregulated pathways include Ribosome and Ribosomal Biogenesis. Notably, among the most downregulated were Peroxisome, Tryptophan metabolism, Citrate cycle, and Propanoate metabolism. The peroxisome participates in the clearance of reactive oxygen species and beta-oxidation of very long fatty acids, while the three later are involved in energy production in mitochondria.

In our search for additional severity biomarkers, we searched for transcripts, clustered by biological processes, correlating with normalized μMax (U18/Ctrl) across all strains. We identified 146 genes associated; 124 positively and 22 negatively correlated (Table S4). The analysis revealed positive correlations with KEEG pathways such as Peroxisome, Glycerolipid Metabolism, Citatrate Cycle, and others (Fig 3H). Interestingly, the cellular components enriched were mitochondria envelope and membrane, suggesting a major role for this organelle in determining disease severity (Fig 3I). Among the negative correlated KEGG and cellular component pathway, we identified the Ribosome (Fig 3J-K). This analysis suggests that the differential severities could be related to alterations in the aforementioned pathways and/or organelle and should be further explored in humans.

### *SLC4A5/bor1* as a therapeutic target for NPC

For modifier validation studies in yeast, we prioritized genes fulfilling the following criteria: i) LOF variants co-segregating with resilience to neurological conditions; ii) Predicted LOF variants exclusively present in the resistant yeast strains (WA and SA); iii) Non-essential genes for yeast viability. This refined our list to *SLC4A5* and *SNRNP200*. To select a single gene for subsequent investigations, we prioritized *SLC4A5* due to its more significant linkage study p value (p=6.1E-07) and for its biological function, a sodium bicarbonate cotransporter crucial for pH regulation (Christensen et al., 2018). In contrast, *SNRNP200* codes for the Small Nuclear Ribonucleoprotein U5 Subunit 200 of the splicesosome potentially regulating thousands of transcripts, making it more difficult to explore its mechanism of action (Wood et al., 2021).

To validate *SLC4A5* as a modifier, we assessed cellular fitness in wild-type (WT; BY4742 strain) and *bor1*-deletant (Δ*bor1*) yeast (derived from the BY4742 background) under control and U18 treatment (200 µg/mL). Growth curves (Figure 4A) were analyzed to determine the maximum specific growth rate (µMax) (Figure 4B). U18-treated Δ*bor1* yeast exhibited a significant increase in μMax compared to its untreated control (fold change 1.68, p=2E-04).

**Figure 4.**
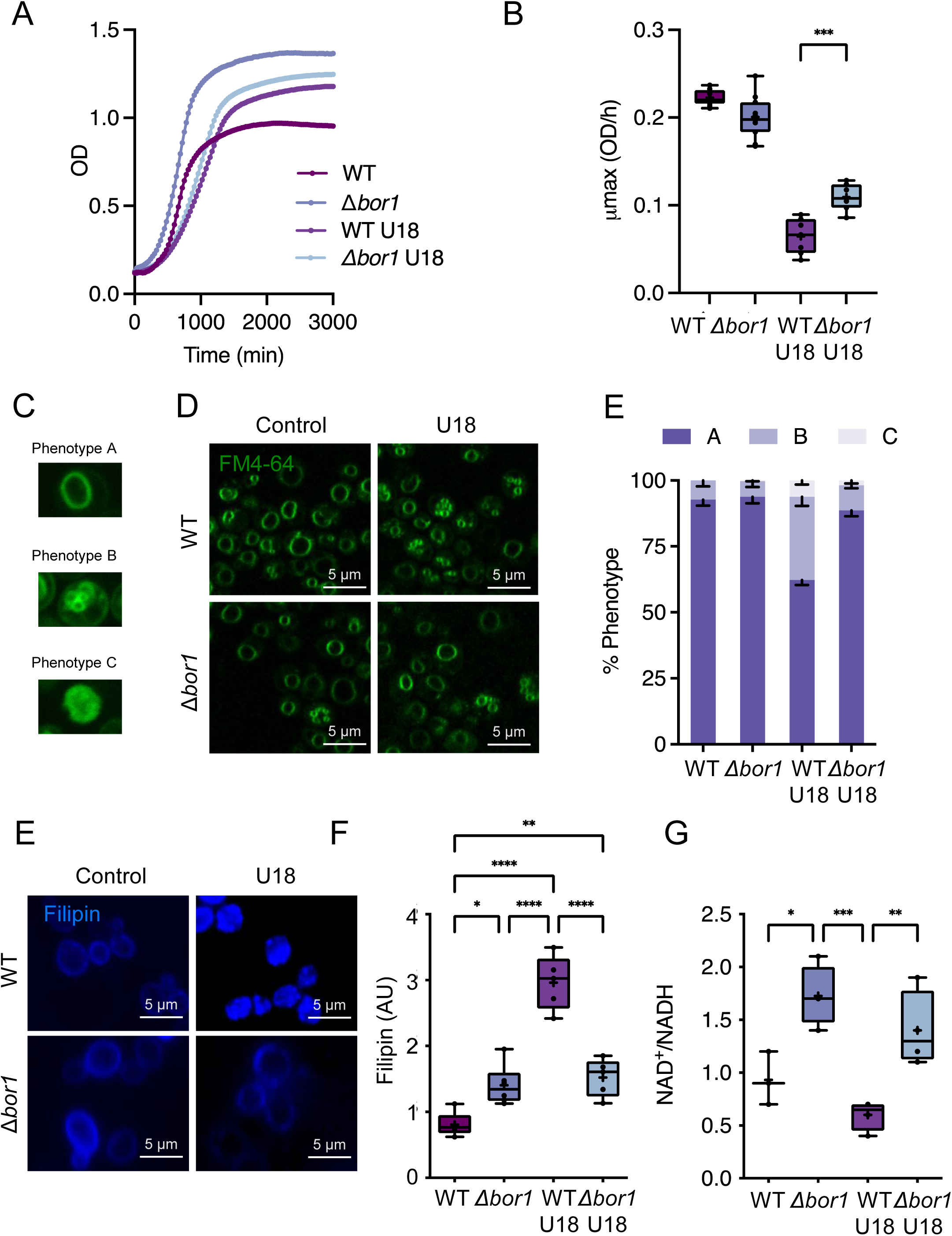
*bor1* as a therapeutic target for NPC-like in yeast. Growth curves of wild-type (WT) and Δ*bor1* yeast, both untreated and after 24 hours of treatment with 200 μg/mL U18. (B) μMax calculated from the growth curves *p < 0.05, **p < 0.01, ***p < 0.001, ****p < 0.0001; Each dot represents a biological replicate (n= 7-12/condition. (C) Examples of vacuolar phenotypes A, B, and C. (D) Representative confocal images of vacuolar morphologies in WT and Δ*bor1* yeast, both untreated and after 24-h U18 treatment. Images were captured using confocal microscopy at 63x magnification. (E) Quantification of vacuolar morphologies for WT and Δ*bor1* yeast under untreated and U18-treated conditions (n=3). (F) Representative images of filipin staining (sterol buildup) in WT and Δ*bor1* yeast strains, both untreated and after 24 hours of 200 μg/mL U18 treatment. (G) Quantification of filipin fluorescence intensity in arbitrary units (A.U.) WT and Δ*bor1* yeast, both untreated and after 24 hours of 200 μg/mL U18 treatment. (F) NAD/NADH ration in WT and Δ*bor1* yeast under control and U18-treated conditions. Each dot represents a biological replicate (n=3-5/condition). Values are represented as mean ± SEM. *p < 0.05, **p < 0.01, ***p < 0.001, **p < 0.0001. Scale bars correspond to 5 mm.

Subsequently, we examined vacuolar morphology in WT and Δ*bor1* yeast under control and after 24 hours of U18 treatment. Vacuoles were categorized into morphologies “A,” “B,” and “C” as previously described (Seeley et al., 2002). Briefly, phenotype A corresponds to one or two large vacuoles. Phenotype B to several well-defined smaller vacuoles and phenotype C to several small, disaggregated vacuoles which are visualized as dots and diffuse staining (Fig 4C-D). Interestingly, Δ*bor1* U18-treated yeast presented a 68% reduction in vacuolar morphology B compared to U18-WT cells (p≤0.0001).

Then, we investigated the impact of *bor1* deletion on sterol accumulation upon NPC-like induction (Figure 4F-G). Filipin staining was employed to quantify ergosterol accumulation in WT and Δ*bor1* yeast, both untreated and following a 24-hour exposure to 200 μg/mL U18. Notably, Δ*bor1* cells exhibited 49% reduction in filipin fluorescence intensity compared to WT under U18 treatment (p≤0.0001, Figure 4F-G). Because we identified associations between mitochondrial transcripts and disease severity, we measured total NAD levels and NAD^+^/NADH ratio. There was no difference in the total NAD levels across WT and Δ*bor1* strains, either under control or U18 exposure (Fig S5). However, the NAD^+^/NADH ratio was different. Δ*bor1* showed 1.88-fold increased levels compared to the WT strain (p<0.05). Moreover, under U18 treatments, Δ*bor1* presented a 2.3-fold increase compared to the WT genotype (p<0.01). Collectively, these findings support the modifier role of *SLC4A5/bor1* for NPC.

## Discussion

Genetics of resilience, defined as genomic factors contributing to maintain functioning in the presence of disease-causing variants, is becoming popular because the discoveries can, potentially, be translated into therapies. Recently, an individual resilient to Alzheimer’s disease (AD), led to the identification of a gain of function variant in *RELN* (Reelin) that prevents neurological symptoms (Lopera et al., 2023). Because for pharmacological purposes it is easier to inhibit a function rather than activated, we focus on uncovering LOF variants that co-segregated with neurological resilience in an NPC family. We successfully used LOF variants in the yeast orthologs to predict disease severity in NPC-like models of diverse genetic backgrounds. Furthermore, transcriptomic analysis in these yeast models linked the expression levels of mitochondrial pathways to cellular fitness. Finally, we validated *SLC4A5/bor1* as a modifier gene and potential therapeutic target for NPC.

Given the substantial phenotypic variability observed among NPC patients, even those with identical mutations, the presence of genetic modifiers is widely recognized (Las Heras et al., 2023). Studies in mouse models with different genetic backgrounds have provided crucial support for this hypothesis. For instance, transferring the *Npc1* mutation from the BALB/c background to the C57BL/6J background in the Pentchev mouse model resulted in markedly different phenotypes (Parra et al., 2011). Furthermore, Rodriguel-Gil et al. conducted a QTL analysis of lifespan in 202 F2 mutant *Npc1* mice derived from crosses between C57BL/6J and BALB/cJ NPC mice, uncovering significant linkage to multiple markers on chromosomes 1 and 7, although specific modifier genes could not be mapped (Rodriguez-Gil et al., 2020). Importantly, the mouse genetic background also influences the response to rapamycin, emphasizing the need for further pharmacogenomic studies in NPC (Calderón & Klein, 2018).

Studies of modifiers in NPC mice have led to the identification of human NPC modifiers. Two sterol O-acyltransferase (SOAT) proteins, SOAT1 and SOAT2, are responsible for converting cholesterol to cholesteryl esters for storage. Genetic deletion of *Soat1* in mouse models of NPC mitigated disease severity, increasing lifespan, while *Soat2* ablation reduced hepatic cholesterol buildup and improved liver function (Lopez et al., 2018; Rogers et al., 2022). Building upon these findings, Farhat et al. investigated the association between a common *SOAT1* variant, rs1044925 (A>C), and disease onset in 117 NPC individuals. Their study revealed a significant association of the C-allele with an earlier age of neurological onset (Farhat et al., 2024).

Identifying biomarkers for disease severity and therapeutic response is challenging and for NPC is an area of active research. Several groups have explored various candidates, including oxysterols, calbindin D, neurofilament light chain, cholestane-triol, amyloid-β, and total and phosphorylated tau, with promising results (Eratne et al., 2024; Gonzalez-Ortiz et al., 2024; Klein et al., 2024; Mengel et al., 2020; Porter et al., 2010; Stern et al., 2024). In a novel approach, van Gool et al. combined neuroimaging studies with plasma proteomic analysis in NPC patients. Their findings revealed enlargements in the choroid plexus in NPC patients, a tissue with high *SLC4A5* expression (Christensen et al., 2013, 2018), associated with elevated plasma IL-18 levels (van Gool et al., 2024). This represents a potentially significant breakthrough as the first identified neuroimaging severity biomarker with a correlated plasma protein, although further clinical validation is necessary. In another study, Baxter et al. conducted a transcriptomic analysis of 41 NPC primary fibroblasts from patients with extensive clinical data. They reported several transcripts associated with disease severity, including *PI4K2A*, a gene that we also found to be correlated with cellular fitness in U18-treated yeast. PI4K2A plays a crucial role in phosphoinositide metabolism, implicated in lysosomal repair and the regulation of lysosomal enzyme activities (Durán et al., 2023; Tan & Finkel, 2022). Recent studies have demonstrated that NPC1 regulates the interaction between the endoplasmic reticulum (ER) and endo-lysosomes, as well as other organelles, controlling the distribution of phosphatidylinositol 4-kinases, including PI4K2A, at lysosomal membranes (Höglinger et al., 2019; Kutchukian et al., 2021). These findings suggest that cells capable of efficiently repairing NPC1-induced lysosomal damage may exhibit faster growth rates, supporting the validity of our approach for identifying biomarkers.

Several lines of evidence point to a strong connection between the NPC pathological cascade and mitochondrial/energy metabolism dysfunction (Torres et al., 2017). NPC cells exhibit elevated mitochondrial cholesterol levels, altered mitochondrial morphology, and increased superoxide production (Balboa et al., 2017). Furthermore, NPC1 inhibition enhances glycolysis while suppressing mitochondrial metabolism in brain microvascular endothelial cells (Moiz et al., 2024). Notably, we identified mitochondrial transcripts and NAD/NADH ratio associated with cellular fitness. This suggests that metabolites-derived from this organelle, including NAD/NADH ratio, represent promising severity biomarkers and that mitochondria could be a therapeutic target for NPC. This aligns with the recent FDA approval of N-Acetyl L-Leucine for NPC treatment, which targets the mitochondrial/energy metabolism pathway (Bremova-Ertl et al., 2024; Moiz et al., 2024). Other molecules targeting this pathway are under investigation for NPC therapeutics (Kataura et al., 2024).

In our study, *SLC4A5* (*bor1*), a sodium bicarbonate cotransporter crucial for pH regulation (Christensen et al., 2018), emerged as a modifier of NPC neurological resilience. Expressed in various tissues, including the brain (Pushkin et al., 2000), *SLC4A5* likely plays a role in maintaining lysosomal pH, which is elevated in NPC (Chakraborty et al., 2017; Tharkeshwar et al., 2017; Wheeler et al., 2019). Reduced *SLC4A5/bor1* activity could potentially normalize lysosomal pH, enhancing enzyme activity and facilitating lipid degradation (Wheeler et al., 2019). This could normalize lysosomal-mitochondrial contact sites and energy production. Altogether, our results support that *SLC4A5* may be a therapeutic target for NPC. Notably, *SLC4A5* variants have been linked to altered cholesterol metabolism, as evidenced by its association with gallstone disease (Ma et al., 2022). Interestingly, NPC1 itself has been implicated in gallstone formation (Morales et al., 2010; Yuan et al., 2005), suggesting that these proteins may cooperate, directly or indirectly, in lipid transport.

In conclusion, studying the genetics of resilience in NPC provides valuable insights into disease mechanisms and potential therapeutic avenues. NPC shares neuropathological features with Alzheimer’s disease (AD) (Langerscheidt et al., 2024) and other lysosomal storage diseases (Yañez et al., 2020). Even some AD cases are caused by *NPC1* variants (Lopergolo et al., 2024). We expect that this work may facilitate the development of biomarkers and of *SLC4A5*-based therapeutics for NPC and other neurodegenerative disorders. In conclusion, by identifying individuals with exceptional resilience, we can uncover disease biomarkers and genetic and molecular pathways that mitigate disease progression.

## Data Availability

RNA-seq data produced are available online at Gene Expression Omnibus (GEO) database, accession no GSE286511.

https://www.ncbi.nlm.nih.gov/geo/query/acc.cgi?acc=GSE286511

## Acknowledgements

We thank Marcelo Rojas-Herrera for technical support. Whole exome sequence (WES) data on the samples analyzed in this study was generated by Heiko Runz, Miriam Stampfer and Jonathon Blake in collaboration with the EMBL Genomics Core facility. Costs for WES were in part covered by Actelion Pharmaceuticals. We also acknowledge the International Niemann-Pick Disease Registry (iNPDR), a European funded project that established a global database for clinical data on patients with Niemann-Pick diseases.

## Author Contributions

M.L.H performed most experiments and wrote the manuscript. B.Z analyzed the RNA-seq. B.Z, V.Ol, J.C.R participated in the yeast experiments. H.R. performed the exome analysis. Y.J. and J.F.C participate in the human linkage analysis. S.Z, E.B. and A.D participate in the experimental design. A.D.K. participated in experimental design, supervised students, funded most of the project, and wrote the manuscript. All the authors edited the text.

## Informed Consent Statement

Informed consent was taken at the University Hospital of Udine, Italy.

## Graphical Abstract

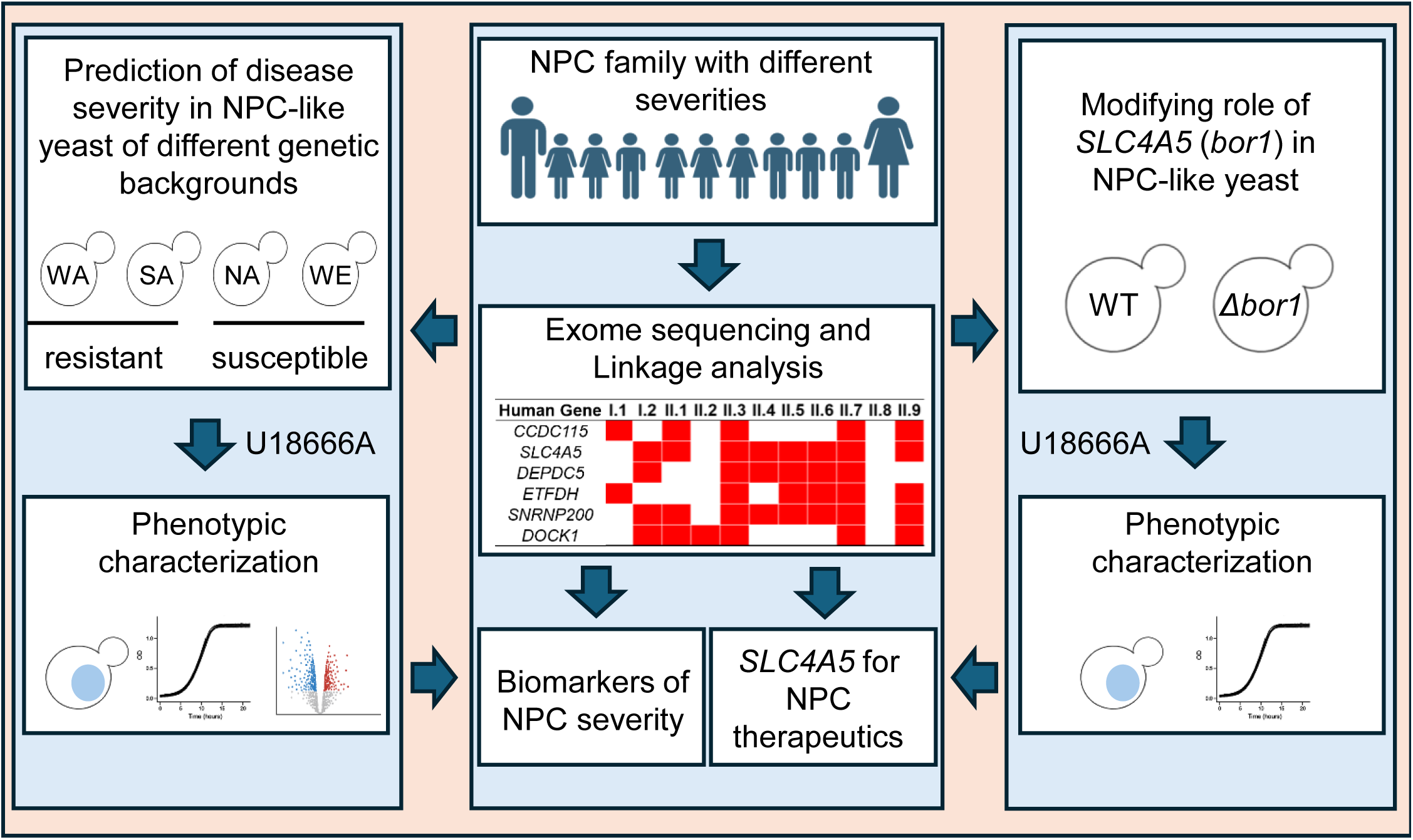

