## Supplementary figures and images for "Genomic Modifiers of Neurological Resilience in a Niemann-Pick C family"

### Supplementary Figure 1

KEGG pathways

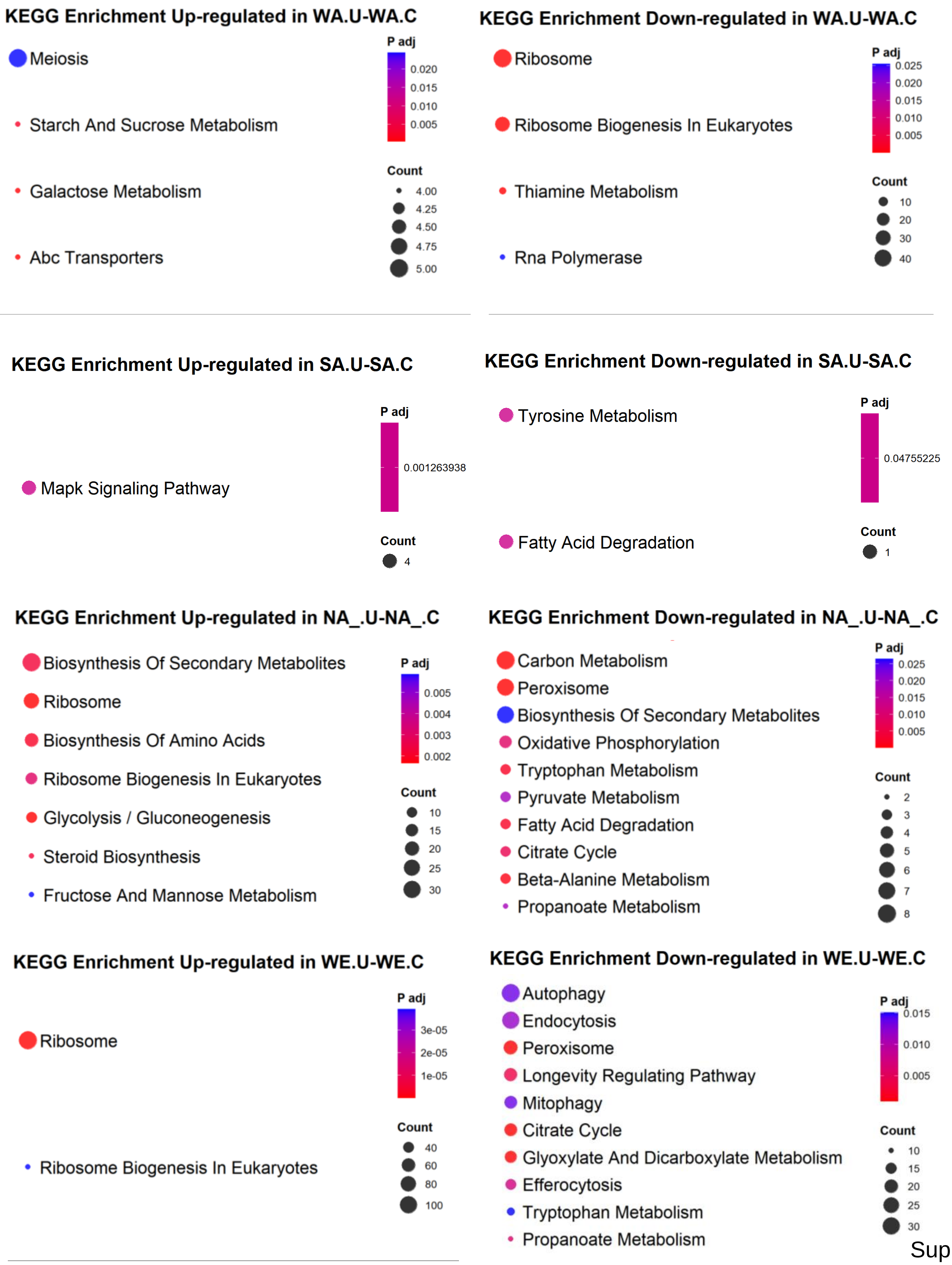


Supp Fig 1

### Supplementary Figure 2

#### Cellular components

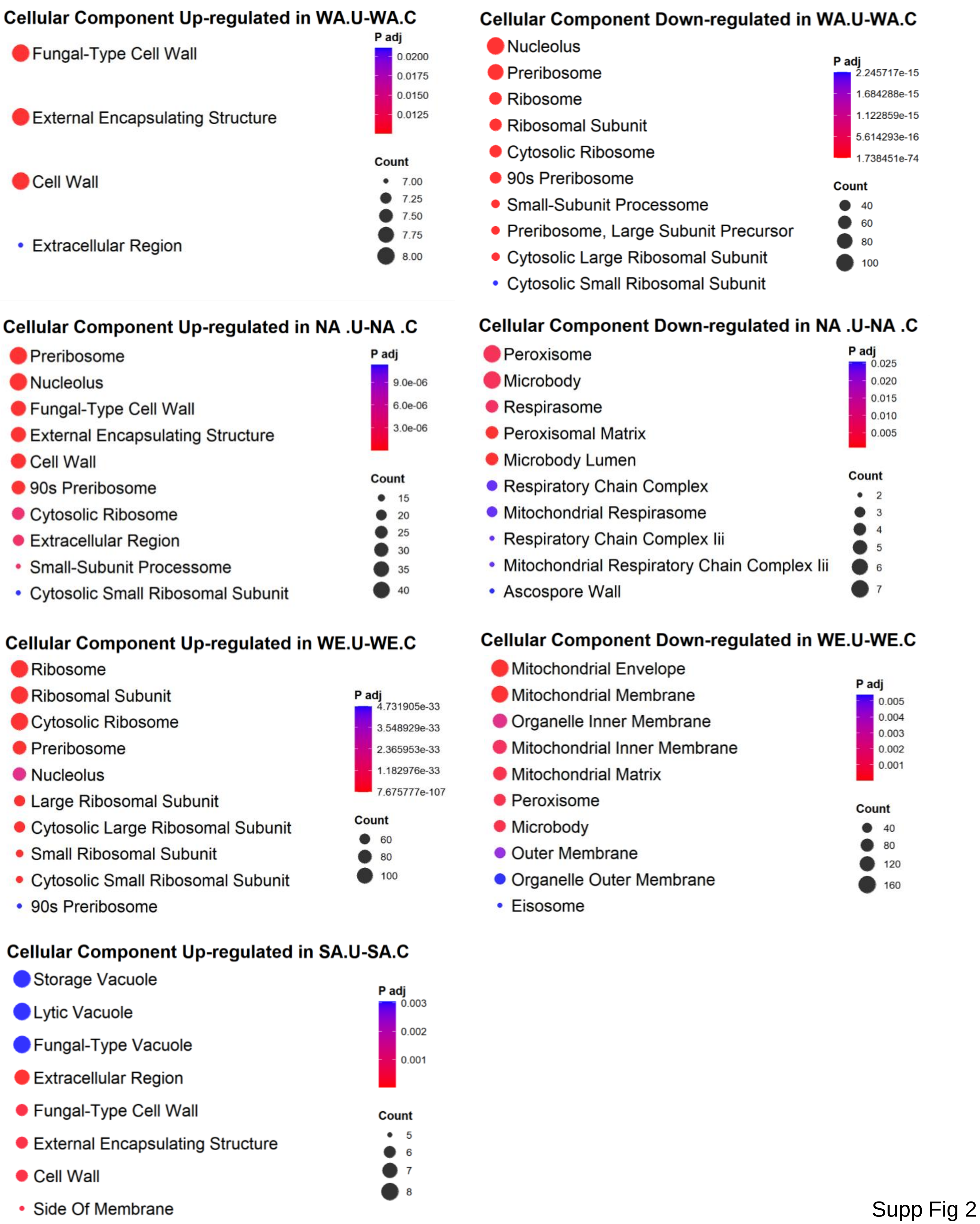


Supp Fig 2

### Supplementary Figure 3

#### Molecular Function

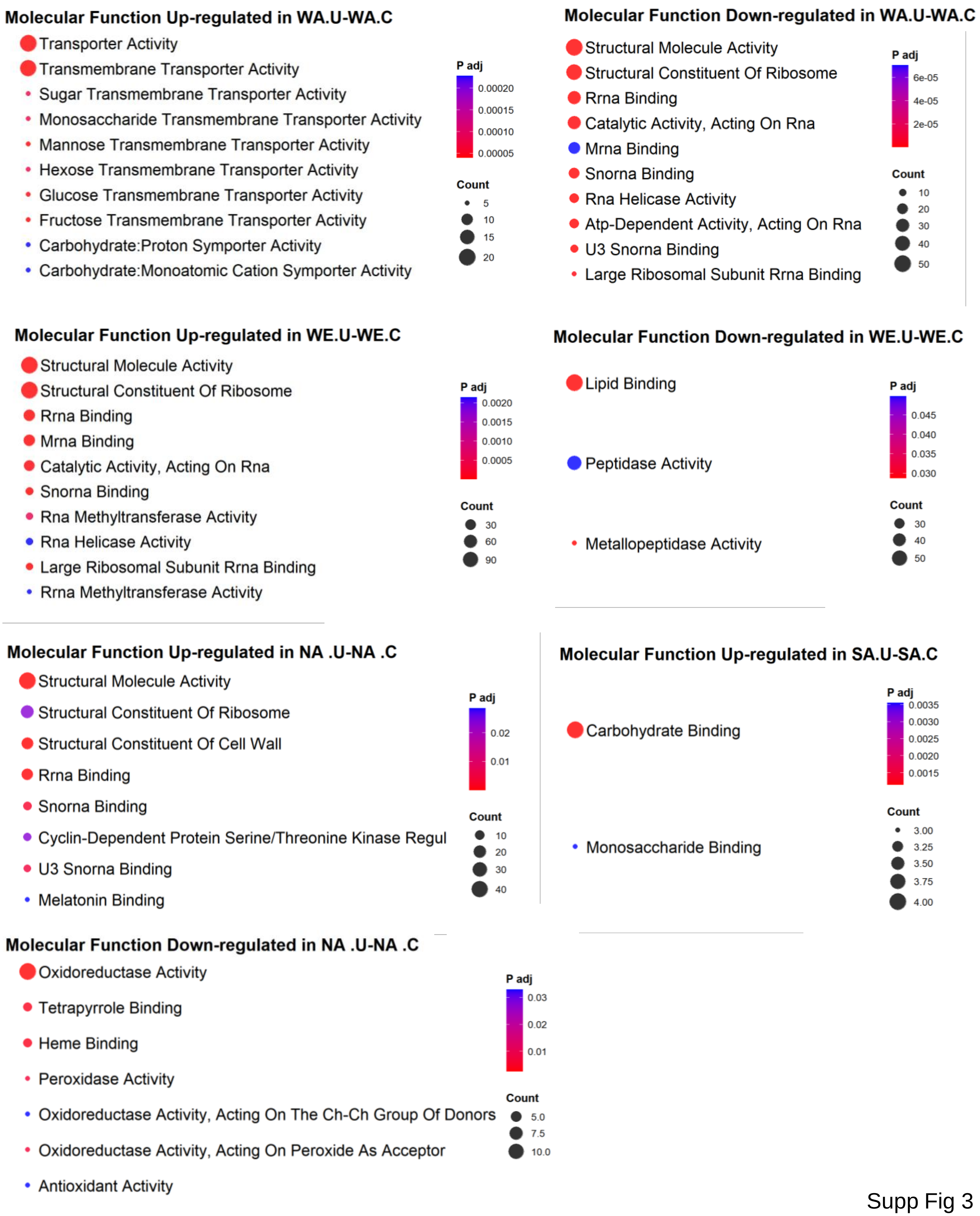


Supp Fig 3

### Supplementary Figure 4

#### Biological Processes

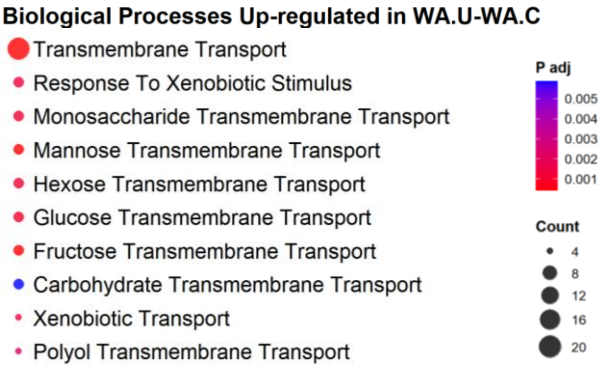


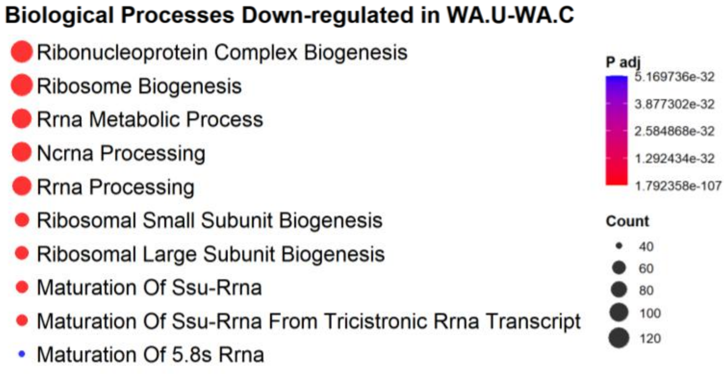


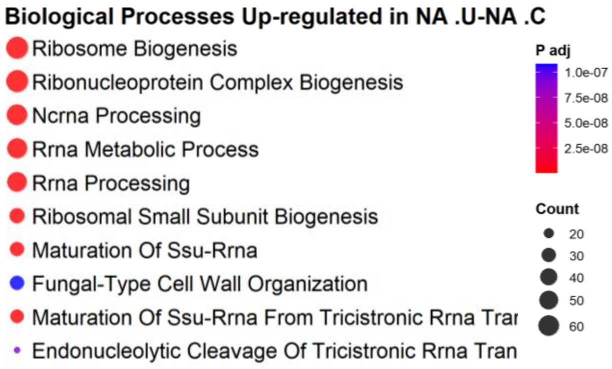


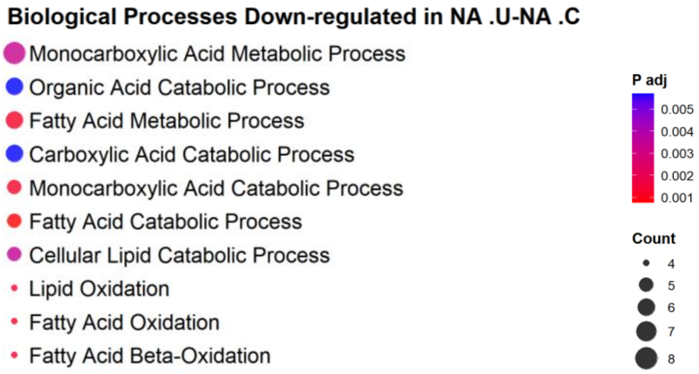


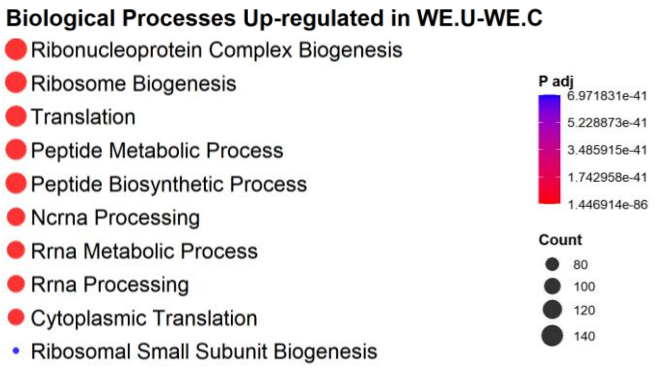


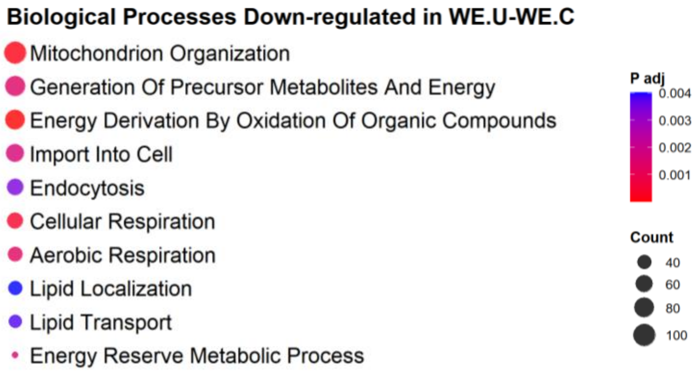


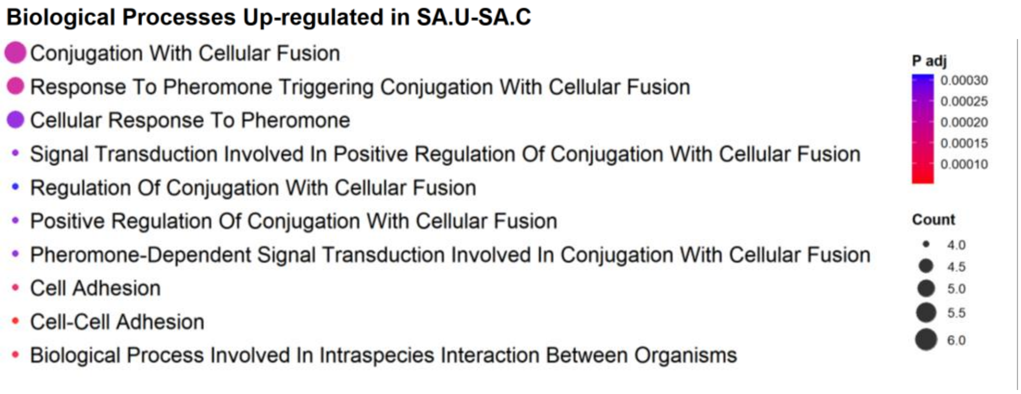


Supp Fig 4

### Supplementary Figure 5

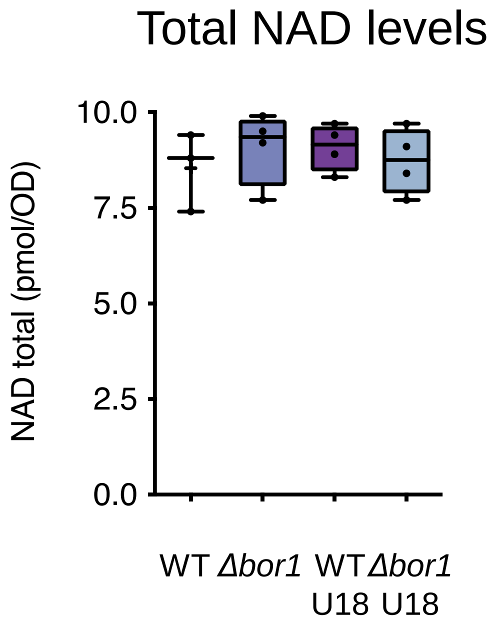
